# Research on Public Risk Perception and Panic Psychology in the Post-Emergency Phase of Major Public Health Crises

**DOI:** 10.1101/2025.04.29.25325326

**Authors:** Ge Shi, Xingtong Men, Jiayi Li, Chenzi Huang, Liyi Ma, Chenchen Xia

## Abstract

This study conducted a questionnaire survey among 1,605 citizens across 32 provinces in China to examine public risk perception and panic psychology regarding COVID-19 pandemic information. A predictive model of risk cognition and psychological behaviors was established to analyze public risk perception characteristics and behavioral responses during the late stages of the pandemic. Findings revealed that government preventive measures had the strongest impact on public risk perception, significantly surpassing the effects of infection-related information, recovery-related data, and personally relevant information. Compared to the 2020 risk perception map, dimensions such as “cure rates” and “post-recovery contagiousness” shifted from unfamiliar and uncontrollable to familiar and controllable, indicating substantial improvements in public risk cognition. While public risk awareness and fear levels decreased significantly compared to the initial pandemic phase, perceived control over “post-recovery health impacts” remained insufficient. Empirical analysis demonstrated that both positive and negative pandemic information could elevate public risk perception, which subsequently influenced psychological behaviors through the mediating role of risk cognition. Furthermore, the moderating effect of psychological tension underwent significant changes as the pandemic evolved, highlighting the dynamic adaptability of public socio-psychological and behavioral patterns during major public health crises. This study provides empirical evidence and scientific insights for risk communication strategies in the post-pandemic era.

## 1 Introduction

As a Major Public Health Emergency, Coronavirus Disease 2019 (COVID-19) has been acknowledged as the most formidable global public health crisis since World War II following its outbreak in late 2019 [1]. Rapidly escalating into a worldwide pandemic, it triggered heightened vigilance across nations. Emerging as a large-scale public health crisis in the social media era with universal human impact, COVID-19 inherently carried substantial risk factors from its inception [2]. As of December 23, 2022, the global cumulative confirmed COVID-19 cases had reached 651.81 million, with fatalities exceeding 6.65 million^1^. Due to the COVID-19 virus’s frequent mutations, high concealment, and rapid transmission, the pandemic has experienced recurrent resurgences. Coupled with the continuously expanding infected population, COVID-19 has inflicted severe negative impacts on individuals, families, and societies. This crisis has posed unprecedented challenges to nations worldwide.

During the pandemic’s progression, public mentality exhibited distinct stage-specific characteristics [3]. Concurrently, researchers have increasingly focused on COVID-19’s psychological impacts. According to World Health Organization (WHO) statistics, the global prevalence of anxiety and depression rose significantly during the pandemic: anxiety cases increased by approximately 25% (from 298 million pre-pandemic to 374 million), while depression cases surged by 28% (from 193 million to 246 million). Furthermore, global prevalence rates for depression and anxiety climbed from 4.4% and 3.6% pre-pandemic to 5.5% and 4.5%, respectively. Studies suggest that beyond the direct traumatic effects of COVID-19, numerous pandemic-related side effects correlate with diminished mental health outcomes [4]. Risk perception—defined as an individual’s subjective interpretation and evaluation of external risks [5]—represents a personal judgment of hazards associated with risk sources [6]. During the early stages of COVID-19, escalating infection rates and intensified containment measures precipitated a severe crisis of trust in society. Misinformation proliferated, public anxiety and panic surged, perceived risks were amplified, and irrational behaviors such as stigmatization, group discrimination, and panic buying became widespread [7]. Confronted with the sudden outbreak, individuals displayed diverse psychological states—including distress, optimism, fear, or calm acceptance—shaped by mixed media messaging and uncertainties in risk perception. These psychological responses further triggered behavioral adaptations, such as mask procurement, frequent handwashing and disinfection, reduced mobility, or dissemination of panic-driven rumors. Crucially, public risk perception of the pandemic has been shown to mediate subsequent psychological manifestations and behavioral coping strategies [8]. Consequently, the role of epidemic information in shaping pandemic trajectories has emerged as a critical research focus [9].

Epidemic Information, as a core element in constructing risk perception during public health crises, serves a dual role: it can heighten public vigilance and motivate proactive protective behaviors, yet simultaneously exacerbate panic, trigger irrational resource depletion, and destabilize societal order [10–13]. From a psychological stress-response perspective, the protracted COVID-19 pandemic constitutes a colossal stressor. Given its high transmissibility and potential to induce multifaceted severe consequences, public risk perception of COVID-19 fundamentally diverges from that of conventional stressors [14]. While risk perception is traditionally assumed to rely on rational judgment, this paradigm faces mounting empirical challenges. Nobel laureate Herbert Simon’s theory of bounded rationality first identified inherent cognitive limitations in human attention and information-processing capacity, which critically constrain objective risk assessment and decision-making [15]. Notably, Daniel Kahneman, another Nobel-winning cognitive psychologist, demonstrated through behavioral economics that even within the framework of expected utility theory, individuals systematically deviate from rational principles [16]. His prospect theory reveals an asymmetric psychological weighting wherein losses are perceived as far more salient than gains, inevitably inducing systematic cognitive biases. Paul Slovic and colleagues identified analogous biases in public risk perception [17]: for emotionally charged issues, expert knowledge and statistical data often fail to override intuitive misjudgments, indicating that cognitive biases operate not merely at the individual level but manifest as structured collective phenomena. These findings collectively indicate that risk perception is far from a purely rational calculus—it is a psychologically constructed process shaped by multiple interacting factors. However, large-scale field investigations of public risk perception and psychological behaviors under high-risk crisis conditions—particularly within Eastern cultural contexts—remain critically underexplored [18–20]. Therefore, this study aims to investigate public psychological responses and behavioral changes during the COVID-19 pandemic, with the dual objectives of providing empirical evidence on risk perception and emergency behaviors in such major public health crises, while offering actionable insights to support governmental contingency planning and policy formulation.

## 2 Research Objectives and Hypotheses

This study is situated within the context of the COVID-19 pandemic as a major public health emergency. In 2022, multiple regions across China experienced renewed outbreaks (Table 1). Against the backdrop of viral mutations, evolving containment strategies, and dynamic shifts in societal psychological adaptation, public response mechanisms may have undergone structural transformations compared to the initial pandemic phase. While widespread vaccination has attenuated fears of perceived fatality risks, the explosive surge in cases driven by the high transmissibility of the Omicron variant and uncertainties surrounding long COVID may have reconfigured public risk perception. To address these dynamics, the research team conducted a nationwide questionnaire survey across 32 provinces in China from February to April 2022. The investigation aims to empirically analyze emerging characteristics of public risk perception and behavioral decision-making, thereby elucidating the dynamic adaptability of socio-psychological and behavioral patterns during protracted public health crises. These findings are intended to provide scientific foundations for risk communication strategies in the post-pandemic era.

**Table 1.**
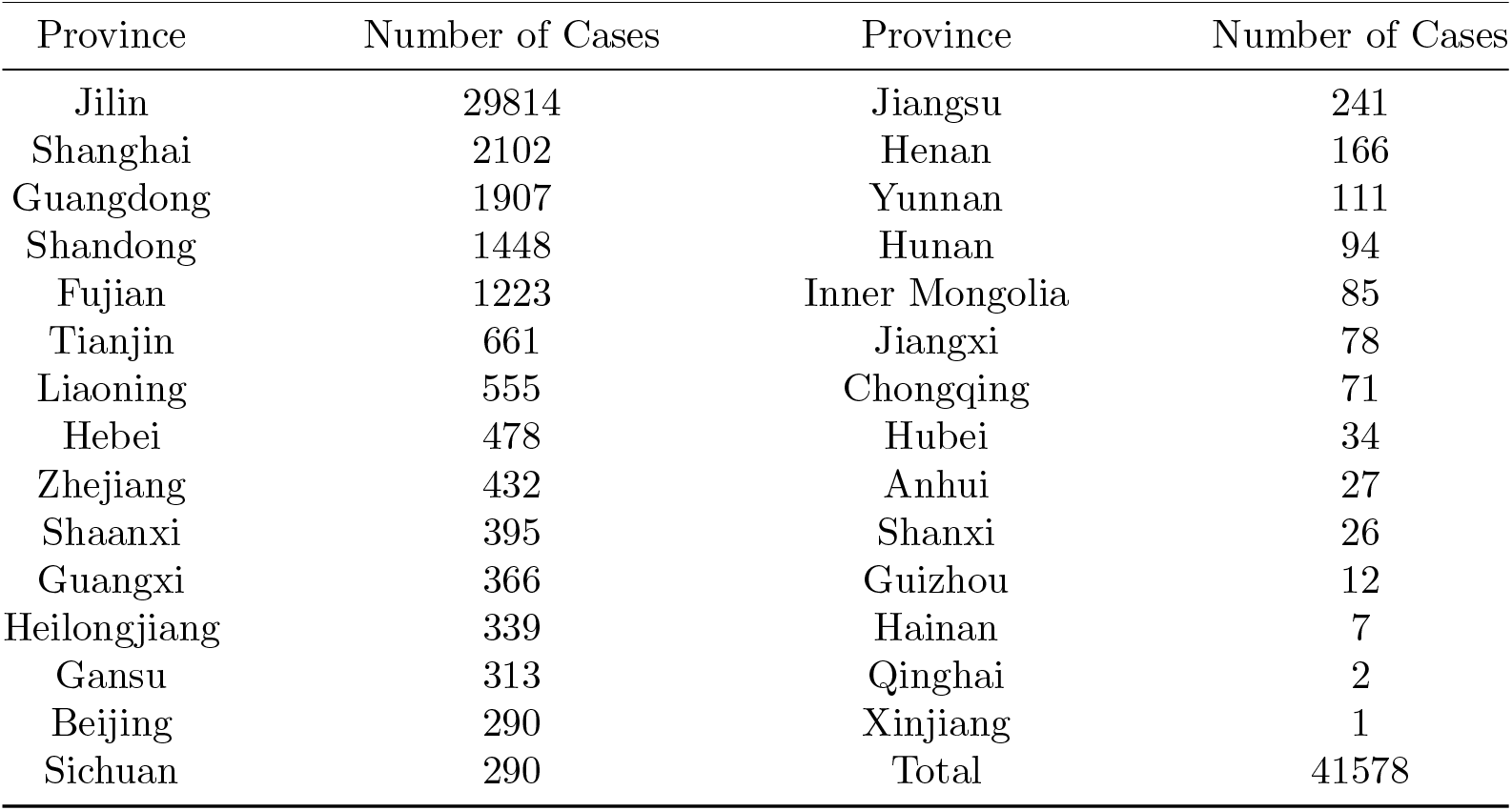
Statistics on Newly Confirmed Cases Across China in March 2022.

| Province | Number of Cases | Province | Number of Cases |
| --- | --- | --- | --- |
| Jilin | 29814 | Jiangsu | 241 |
| Shanghai | 2102 | Henan | 166 |
| Guangdong | 1907 | Yunnan | 111 |
| Shandong | 1448 | Hunan | 94 |
| Fujian | 1223 | Inner Mongolia | 85 |
| Tianjin | 661 | Jiangxi | 78 |
| Liaoning | 555 | Chongqing | 71 |
| Hebei | 478 | Hubei | 34 |
| Zhejiang | 432 | Anhui | 27 |
| Shaanxi | 395 | Shanxi | 26 |
| Guangxi | 366 | Guizhou | 12 |
| Heilongjiang | 339 | Hainan | 7 |
| Gansu | 313 | Qinghai | 2 |
| Beijing | 290 | Xinjiang | 1 |
| Sichuan | 290 | Total | 41578 |

This study seeks to validate the following hypotheses:

1. The unknown factors and uncontrollable elements inherent to the COVID-19 pandemic are critical determinants shaping public risk perception.
2. Compared to the initial pandemic phase, public risk awareness and perceived threat severity exhibit significant mitigation in later stages of the COVID-19 crisis, reflecting adaptive psychological responses to prolonged exposure.
3. Both positive and negative pandemic-related information may elevate public risk perception levels.
4. COVID-19-related information influences psychological and behavioral outcomes through the mediating factor of risk perception, with predictive indicators including psychological tension, coping behaviors, and mental health status.
5. “Personally relevant information” represents a unique category that not only indirectly promotes proactive coping behaviors via risk perception mediation but also directly triggers avoidant coping strategies.

## 3 Research Methods

### 3.1 Survey Method and Questionnaire Design

Based on existing literature and aligned with the research objectives, this study designed a questionnaire to collect data on public risk perception during the COVID-19 pandemic. The questionnaire was administered and collected through Credamo (https://www.credamo.com), a publicly accessible intelligent data acquisition platform. Prior to distribution, the platform explicitly informed participants of the research objectives, data utilization purposes, privacy protection measures, and withdrawal mechanisms via pop-up notifications and user agreements, ensuring autonomous decision-making based on comprehensive understanding. Consequently, all participants in this study engaged voluntarily through the platform. The platform strictly adheres to data privacy protection regulations and incorporates an ethical risk assessment module, requiring researchers to declare data usage intentions, potential risks, and corresponding mitigation measures during questionnaire submission. This framework satisfies dual safeguards for participant voluntariness and ethical compliance.Empirical research methods were applied to further validate the data. The questionnaire covered five sections:

1. Perception of pandemic information
2. Risk cognition assessment
3. Psychological states
4. Coping behaviors
5. Psychological indicators

Variables were designed by referencing prior scholarly findings [21] and integrating them with the specific context of the COVID-19 pandemic. Details are as follows:

Epidemic Information Perception Questionnaire: Existing research in risk communication categorizes risk information into two components: information about the risk itself and information about measures to mitigate the risk [22]. Accordingly, this study divides information factors influencing risk perception into two parts: COVID-19-related information (e.g., disease characteristics, transmissibility, mortality rate) and preventive measures (e.g., government directives, containment policies). The questionnaire items were designed based on prior studies by Shi Kan et al., totaling 23 items. A 5-point Likert scale was used, ranging from 1 = no impact to 5 = significant impact.

Risk Perception Assessment Questionnaire: Drawing on Slovic’ s risk perception model [18], this study measured public risk cognition using two dimensions: familiarity and controllability. Assessed risk events included: COVID-19 etiology, transmission routes, cure rates, preventive measures, post-recovery health impacts, post-recovery contagiousness, and overall perception of coronaviruses. Both dimensions employed 5-point Likert scales: familiarity ranged from 1 = very unfamiliar to 5 = very familiar, and controllability from 1 = completely uncontrollable to 5 = fully controllable.

Mental Health Evaluation Questionnaire: Adapted from Goldberg’ s (1992) GHQ-12 items [21], this section included statements such as ability to concentrate on tasks, feeling worthless, and insomnia due to anxiety. Responses were measured on a 5-point Likert scale from 1 = never to 5 = always.

Coping Behavior Questionnaire: Based on Moos’ s (1993) Coping Response Inventory [23], this questionnaire comprised 8 items across two dimensions: proactive coping (e.g., emphasizing disinfection and handwashing habits) and avoidant coping (e.g., reducing social interactions). Proactive coping was further divided into self-protection and active problem-solving.

Psychological Indicators Questionnaire: This questionnaire included 4 items assessing risk perception intensity and psychological tension, measured using a 5-point Likert scale.

### 3.2 Sample Selection and Data Collection

Studies indicate that risk perception and anxiety levels among populations in areas with varying pandemic severity exhibit a “ripple effect”, with higher levels observed in severely affected regions compared to others [24]. Therefore, in February 2022, the research team employed a stratified sampling method to distribute questionnaires across 32 provinces (excluding Hong Kong and Macao) in China, aiming to comprehensively assess residents’ risk perception across regions with differing outbreak intensities.

Existing research also suggests that respondents with lower education levels and higher monthly incomes tend to overestimate or underestimate risks, reporting stronger positive emotional responses to the pandemic, while those with risk underestimation exhibit weaker negative emotional reactions [25]. Additionally, children, older adults, and disadvantaged socioeconomic groups are generally considered more vulnerable to pandemic impacts: children and older adults face higher infection risks, while disadvantaged groups experience greater disruptions to employment and daily life, potentially leading to distinct sociopsychological profiles compared to other demographics [3]. Consequently, the study targeted populations across diverse age groups, educational backgrounds, and occupations.

A total of 1,772 questionnaires were collected. After excluding 167 invalid responses, 1,605 valid samples were retained, yielding a validity rate of 90.41%. The demographic distribution of the survey is presented below:

The sample comprised 1,605 participants, with 37.5% (*n* = 602) male and 62.5% (*n* = 1, 003) female. Age distribution was as follows: 36 participants under 20 years old, 836 aged 20–29 years, 640 aged 30–39 years, 67 aged 40–49 years, 25 aged 50–59 years, and 1 individual over 60 years. In terms of educational attainment, 166 held an associate degree or lower qualifications, 1,172 had a bachelor’s degree, and 267 possessed a master’s degree or higher. Occupational categories included 376 students, 897 corporate employees, 68 civil servants, 56 service industry workers, 17 healthcare workers, 116 professionals in science, education, or cultural fields (e.g., researchers, teachers, artists), 34 self-employed individuals, 20 laborers (blue-collar workers), and 21 individuals in other occupations (e.g., farmers, police officers, military personnel).

### 3.3 Reliability and Validity Tests

Following the survey, reliability and validity analyses were conducted as critical steps to ensure data quality prior to formal analysis. These analyses not only verify the accuracy and consistency of the sample data but also assess whether the questionnaire adequately reflects the research objectives.

**Table 2.** Demographic Distribution of Survey Participants.

| PR | *n* | PR | *n* | Demographics | % | Demographics | % |
| --- | --- | --- | --- | --- | --- | --- | --- |
| Guangdong | 226 | Anhui | 50 | Sex |  | Occupation |  |
| Shandong | 123 | Hunan | 48 | Male | 37.5 | Civil Servant | 4.24 |
| Jiangsu | 115 | Jiangxi | 46 | Female | 62.5 | Corporate Employee | 55.89 |
| Beijing | 103 | Shaanxi | 31 | Age |  | Service Worker | 3.49 |
| Henan | 101 | Chongqing | 30 | < 20 | 2.24 | Healthcare Worker | 1.06 |
| Shanxi | 77 | Jilin | 23 | 20-29 | 52.09 | Laborer | 1.25 |
| Hubei | 70 | Guizhou | 19 | 30-39 | 39.88 | Self-employed | 2.12 |
| Sichuan | 68 | Hainan | 16 | 40-49 | 4.17 | Military Personnel | 0.12 |
| Hebei | 63 | Tianjin | 16 | 50-59 | 1.56 | Student | 23.43 |
| Heilongjiang | 59 | Yunnan | 15 | ≥ 60 | 0.06 | Professional in S/E/C | 7.23 |
| Shanghai | 58 | Neimenggu | 9 | Education |  | Farmer | 0.06 |
| Liaoning | 57 | Xinjiang | 6 | ≤ Junior high | 0.19 | Police Officer | 0.06 |
| Guangxi | 57 | Ningxia | 5 | High/Vocational | 1.99 | Retired Individual | 0.06 |
| Zhejiang | 56 | Gansu | 4 | Associate | 8.16 | Other | 0.69 |
| Fujian | 50 | Qinghai | 1 | Bachelor | 73.02 | Unemployed | 0.31 |
| Total | 1602 |  |  | ≥ Master | 16.64 |  |  |

#### 3.3.1 Reliability Test

Cronbach’s alpha coefficient was used to evaluate the internal consistency (reliability) of the questionnaire. The overall reliability of all items was tested, with results shown in Table 3. The Cronbach’s alpha coefficient was 0.916, exceeding the threshold of 0.9, indicating excellent reliability and high consistency among the questionnaire items. This demonstrates that the questionnaire is well-designed and highly reliable.

**Table 3.** Reliability Analysis Results.

|  | Cronbach's Alpha | Number of Items |
| --- | --- | --- |
| Entire Questionnaire | 0.916 | 71 |

#### 3.3.2 Validity Test

This study conducted validity analysis of the scale questionnaire using SPSS, with results presented in Table 4. The Kaiser-Meyer-Olkin (KMO) measure yielded a value of 0.942 (above 0.9), indicating strong correlations among variables and high suitability for exploratory factor analysis. Furthermore, Bartlett’s Test of Sphericity showed a p-value of 0.000 (below the significance level of 0.05), confirming that the correlation matrix significantly deviated from an identity matrix. These results demonstrate robust validity of the questionnaire data, supporting further analytical procedures.

**Table 4.** Validity Test Results.

|  | KMO & Bartlett's Test |  |
| --- | --- | --- |
|  | KMO | 0.942 |
|  | Approximate Chi-Square | 57165.681 |
| Bartlett | df | 2485 |
|  | p | 0.000 |

## 4 Results and Analysis

### 4.1 Analysis of Information Factors Influencing Risk Perception

To determine the optimal number of factors and validate the dimensionality of the questionnaire, a varimax-rotated factor analysis was performed on the 23 information items affecting risk perception. The analysis identified four key factors, with a cumulative variance contribution rate of 60.106% (Table 5), indicating minimal information loss in the original questionnaire and satisfactory factor analysis outcomes.

**Table 5.** Total Variance Explained.

| Component | Initial Eigenvalues |  |  | Sum of Squared Loadings<br>(Extraction) |  |  | Sum of Squared Loadings<br>(Extraction) |  |  |
| --- | --- | --- | --- | --- | --- | --- | --- | --- | --- |
|  | Total | % of<br>Variance | Cumul<br>-ative % | Total | % of<br>Variance | Cumul<br>-ative % | Total | % of<br>Variance | Cumul<br>-ative % |
| 1 | 7.345 | 31.934 | 31.934 | 7.345 | 31.934 | 31.934 | 4.925 | 21.414 | 21.414 |
| 2 | 3.386 | 14.721 | 46.655 | 3.386 | 14.721 | 46.655 | 4.318 | 18.772 | 40.185 |
| 3 | 1.716 | 7.459 | 54.115 | 1.716 | 7.459 | 54.115 | 2.841 | 12.350 | 52.536 |
| 4 | 1.378 | 5.991 | 60.106 | 1.378 | 5.991 | 60.106 | 1.741 | 7.570 | 60.106 |
| 5 | 0.749 | 3.258 | 63.364 |  |  |  |  |  |  |
| 6 | 0.684 | 2.974 | 66.338 |  |  |  |  |  |  |
| 7 | 0.642 | 2.791 | 69.129 |  |  |  |  |  |  |
| 8 | 0.638 | 2.773 | 71.902 |  |  |  |  |  |  |
| 9 | 0.617 | 2.681 | 74.584 |  |  |  |  |  |  |
| 10 | 0.572 | 2.485 | 77.069 |  |  |  |  |  |  |
| 11 | 0.545 | 2.371 | 79.439 |  |  |  |  |  |  |
| 12 | 0.531 | 2.308 | 81.748 |  |  |  |  |  |  |
| 13 | 0.508 | 2.210 | 83.957 |  |  |  |  |  |  |
| 14 | 0.478 | 2.077 | 86.034 |  |  |  |  |  |  |
| 15 | 0.450 | 1.954 | 87.988 |  |  |  |  |  |  |
| 16 | 0.426 | 1.851 | 89.839 |  |  |  |  |  |  |
| 17 | 0.418 | 1.817 | 91.656 |  |  |  |  |  |  |
| 18 | 0.405 | 1.760 | 93.416 |  |  |  |  |  |  |
| 19 | 0.381 | 1.657 | 95.073 |  |  |  |  |  |  |
| 20 | 0.349 | 1.517 | 96.590 |  |  |  |  |  |  |
| 21 | 0.297 | 1.293 | 97.883 |  |  |  |  |  |  |
| 22 | 0.253 | 1.101 | 98.984 |  |  |  |  |  |  |
| 23 | 0.234 | 1.016 | 100.000 |  |  |  |  |  |  |

Extraction Method: Principal Component Analysis.

To visually and accurately determine the optimal number of factors, a scree plot was generated for this study. As shown in Figure 1, the eigenvalues level off after extracting four components, indicating a significant reduction in marginal explanatory power beyond this point. The study thus concludes that pandemic-related information can be categorized into four distinct dimensions for deeper analysis.

**Fig 1.**
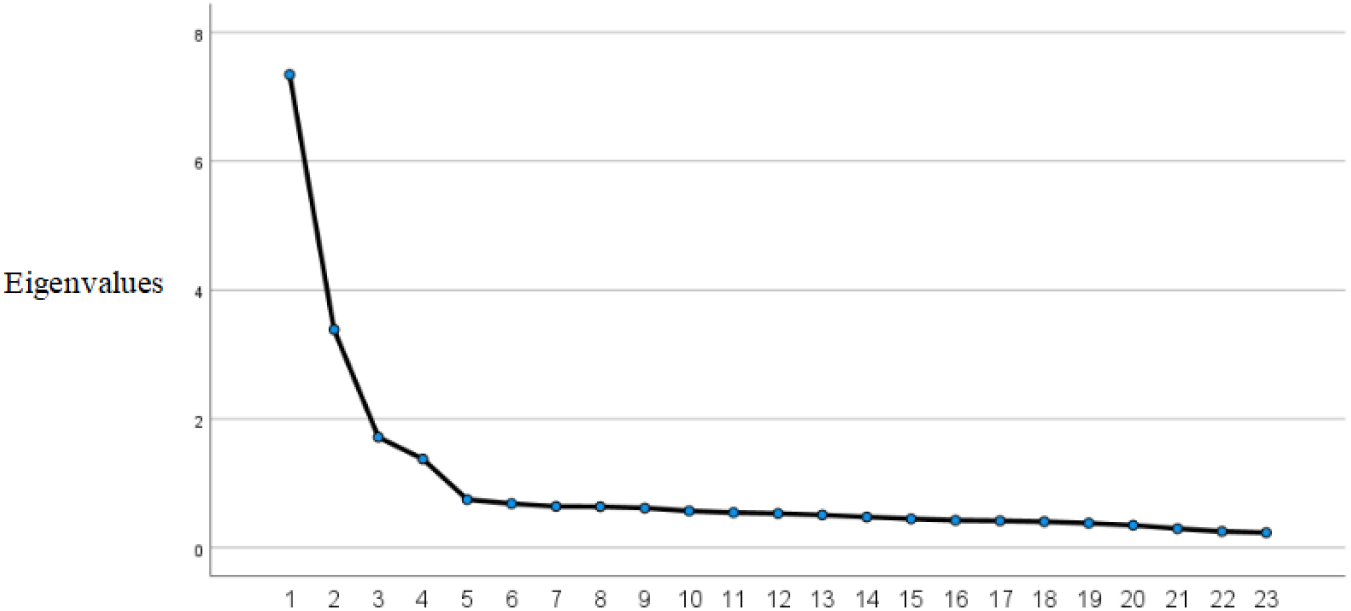
Scree Plot of Variance

While the scree plot and total variance matrix alone could not definitively identify variables with the highest contributions, the study further examined the composition of the four key factors through the rotated component matrix generated by SPSS (Table 6). Factor A, termed “Infection-Related Information”, encompasses eight negative pandemic indicators such as cumulative confirmed cases, quarantined individuals, and daily new infections. As a core element of public health governance, infection-related information not only shapes collective cognition but also profoundly impacts socio-psychological stability, with its scientific validity and dissemination modes directly influencing individual risk assessments. Factor B, “Recovery-Related Information”, includes two positive indicators: daily recoveries and discharged patients. From a governance perspective, recovery data serves dual roles as a risk buffer and behavioral guide. Scientifically reported recovery rates mitigate fear induced by infection statistics, while survivor tracking enhances disease outcome predictability, reframing societal risk perception. This cognitive restructuring aligns with Slovic’s risk perception model, wherein improved recovery rates reduce perceived uncontrollability, thereby recalibrating individual psychological weighting of pandemic threats. Factor C, “Personally Relevant Information”, comprises four negative indicators reflecting direct exposure risks within individuals’ geospatial environments (e.g., confirmed cases in workplaces or residential areas) and social networks (e.g., infected acquaintances). These metrics assess viral penetration in physical spaces and relational networks, triggering reevaluation of daily risk exposure and social behavior safety. Factor D, “Government Preventive Measures”, integrates nine positive items such as treatment/environmental improvement reports, transmission-blocking policies, and travel advisories. Multifaceted interventions shape public risk perception and behavioral choices through mechanisms like credibility halo effects from high-frequency official communications (e.g., press briefings), which lower information-screening costs and promote adoption of government-endorsed risk assessments. Simultaneously, risk cognition emerges from dual pathways: interpersonal trust transmission (e.g., pandemic advice from acquaintances, which shows a 4.7× higher adoption rate than from unknown sources) and public information diffusion (e.g., social media platforms, where information overload may induce cognitive paralysis or polarized decisions). Empirical findings further reveal that anxiety propagation within close networks amplifies chain emotional contagion.

**Table 6.** Rotated Component Matrix.

|  | A | B | C | D |
| --- | --- | --- | --- | --- |
| 1(A1) | 0.718 |  |  |  |
| 2(A2) | 0.698 |  |  |  |
| 3(A3) | 0.654 |  |  |  |
| 4(A4) | 0.698 |  |  |  |
| 5(A5) | 0.783 |  |  |  |
| 6(B1) |  | 0.886 |  |  |
| 7(B2) |  | 0.881 |  |  |
| 8(A6) | 0.756 |  |  |  |
| 9(A7) | 0.705 |  |  |  |
| 10(A8) | 0.735 |  |  |  |
| 11(C1) |  |  | 0.825 |  |
| 12(C2) |  |  | 0.823 |  |
| 13(C3) |  |  | 0.799 |  |
| 14(C4) |  |  | 0.733 |  |
| 15(D1) |  |  |  | 0.718 |
| 16(D2) |  |  |  | 0.652 |
| 17(D3) |  |  |  | 0.697 |
| 18(D4) |  |  |  | 0.744 |
| 19(D5) |  |  |  | 0.707 |
| 20(D6) |  |  |  | 0.743 |
| 21(D7) |  |  |  | 0.685 |
| 22(D8) |  |  |  | 0.671 |
| 23(D9) |  |  |  | 0.732 |

The study concludes that these four factors—validated through rotated component analysis and consistent with prior research by Shi Kan et al.—constitute the most critical dimensions shaping public risk perception during the COVID-19 pandemic.

The analysis of variance (ANOVA) and descriptive statistics revealed significant differences in the impact levels among the four information factors, with pairwise comparisons showing distinct variations. Matrix analysis identified “personally relevant information” as the most influential factor (mean impact score = 3.65), followed by “government preventive measures” (3.61), “infection-related information” (3.60), and “recovery-related information” (3.07). This hierarchy suggests a public tendency to prioritize negative information during pandemics, attributable to heightened neural processing of threat-related stimuli under stress [26]. Furthermore, optimistic bias theory posits that individuals systematically underestimate personal susceptibility to risks, a psychological defense mechanism that diminishes the regulatory effect of recovery-related information on risk perception.

Further examination of key risk assessment drivers identified the most impactful indicators within each factor. For Factor A (infection-related information), “daily new infections” exerted the strongest influence (3.67), categorized between moderate and high impact. In Factor B (recovery-related information), “daily recoveries” showed the highest effect (3.07), similarly within the moderate-to-high range. Factor C (personally relevant information) was most strongly affected by “presence of confirmed cases in workplaces or residential areas” (3.81), while Factor D (government preventive measures) demonstrated peak sensitivity to “transmission-blocking measures” (3.65), both scores indicating moderate-to-high influence thresholds.

### 4.2 Analysis of Public Risk Perception Characteristics

#### 4.2.1 Analysis of Risk Information Cognition

As shown in Table 7, ANOVA results revealed significant variations in public familiarity with risk events. The familiarity levels (from lowest to highest) were ranked as follows: post-recovery health impacts, post-recovery contagiousness, coronavirus etiology, cure rates, preventive measures and efficacy, and transmission routes and infectivity.

**Table 7.** Public Risk Perception Statistics.

| Risk Event | Familiarity |  | Controllability |  |
| --- | --- | --- | --- | --- |
|  | M | SD | M | SD |
| 1.Etiology of Coronavirus | 3.43 | 0.809 | 3.14 | 0.772 |
| 2.Transmission Routes and Infectivity | 3.71 | 0.963 | 3.37 | 0.820 |
| 3.Cure Rate | 3.46 | 0.796 | 3.40 | 0.739 |
| 4.Preventive Measures and Efficacy | 3.68 | 0.926 | 3.57 | 0.827 |
| 5.Post-Recovery Health Impacts | 3.26 | 0.824 | 3.04 | 0.760 |
| 6.Post-Recovery Contagiousness | 3.33 | 0.889 | 3.24 | 0.785 |
| 7.Overall Perception of COVID-19 | 3.55 | 0.830 | 3.36 | 0.735 |

Similarly, public perceived controllability of risk events also exhibited heterogeneity. Controllability rankings (from lowest to highest) were: post-recovery health impacts, etiology, post-recovery contagiousness, transmission routes and infectivity, cure rates, and preventive measures and efficacy.

#### 4.2.2 Comparative Analysis of Risk Perception Distribution Across Two Pandemic Outbreak Periods

Based on the findings, a risk perception spatial distribution map for public COVID-19 risk cognition was developed (Figure 2), displaying item numbers for clarity. This map was then compared to the 2020 pandemic risk perception map (Figure 3) published by Shi Kan et al. to identify temporal shifts in public cognitive patterns^2^.

**Fig 2.**
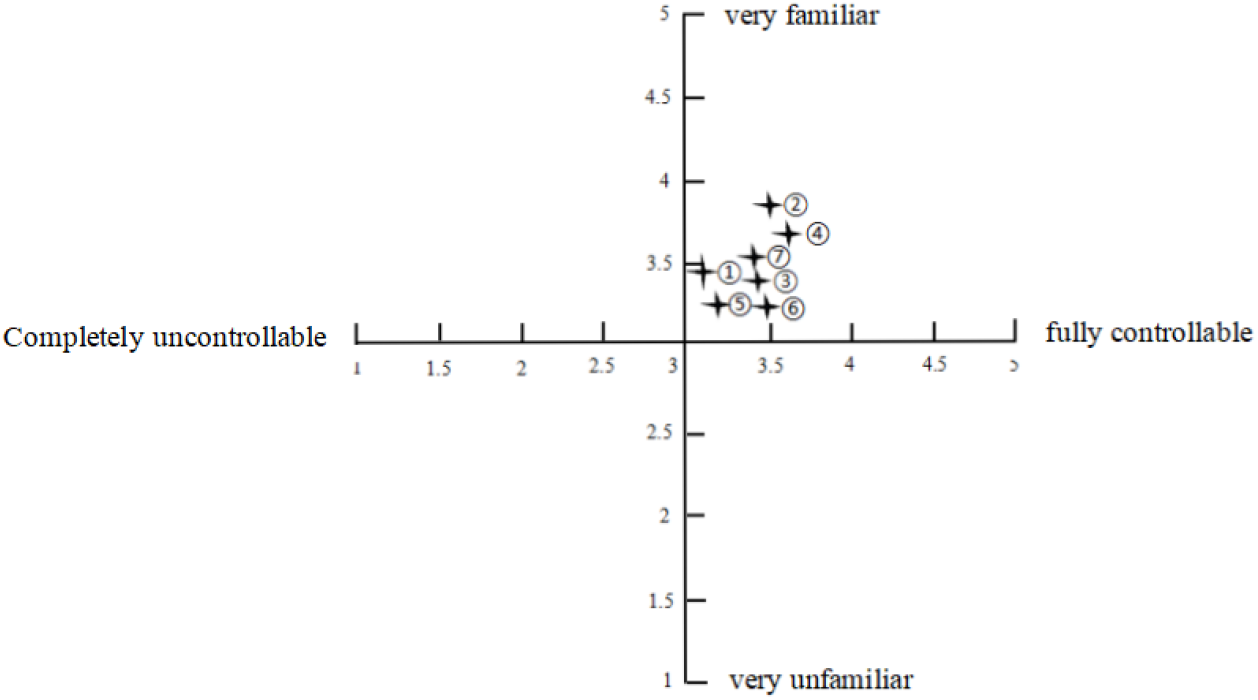
Spatial Distribution of Public Risk Perception Characteristics Regarding COVID-19 in 2022

**Fig 3.**
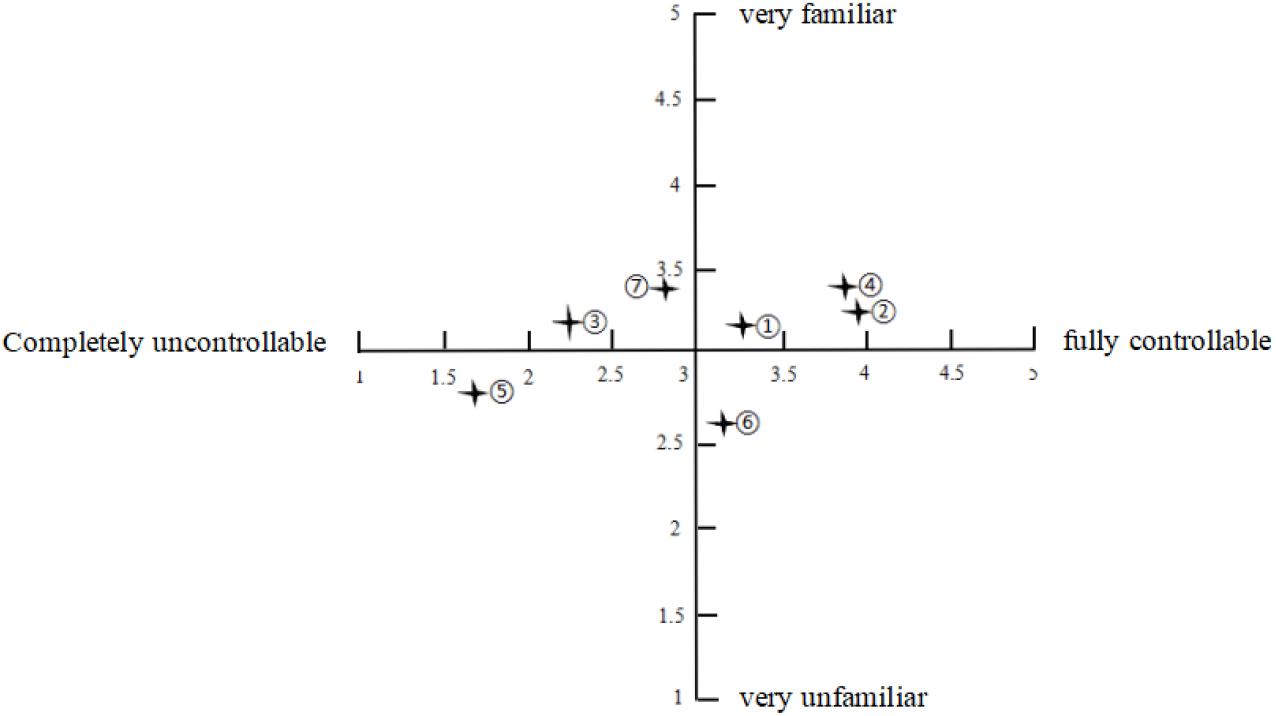
Risk Perception Map of Pandemic Information of 2020 (from Shi Kan et al.)

Comparative analysis of Figures 2 and 3 reveals distinct shifts in public risk perception between the 2020 and 2022 pandemic phases. As illustrated in Figure 2, the aggregate public risk perception of COVID-19 during February-April 2022 clustered toward the upper-right quadrant of the risk factor space, characterized by high familiarity and controllability. This spatial distribution contrasts markedly with the 2020 baseline (Figure 3), reflecting a societal adaptation process shaped by cumulative pandemic experience. Specifically, public familiarity and perceived control over transmission routes, preventive efficacy, cure rates, and post-recovery contagiousness transitioned from moderate (2020) to high levels (2022). These shifts correlate with China’s enhanced medical response capabilities—including reduced mortality rates, diminished clinical severity of viral variants, and optimized containment protocols—as well as improved public health literacy. Notably, post-recovery health impacts and viral etiology remained positioned in low controllability zones, indicating persistent public uncertainty regarding long-term sequelae and evolving viral pathogenesis. Despite systemic advancements, these dimensions retained elevated risk sensitivity, aligning with Hypothesis 1’s proposition of pandemic-specific risk cognition features. Public anxiety toward unresolved biological uncertainties (e.g., unpredictable post-recovery complications, mutational mechanisms of SARS-CoV-2) underscores the cognitive dominance of uncontrollable factors.

The observed partial validation of Hypothesis 2—evidenced by reduced fear toward controllable risks (e.g., transmission mitigation) but sustained apprehension toward health impacts and etiology—highlights critical vulnerabilities for crisis management. Residual gaps in controllability, particularly concerning long-term health consequences and pathogen evolution, may amplify panic during future outbreaks, necessitating targeted risk communication strategies to address these latent cognitive triggers.

### 4.3 Analysis of Public Psychological and Behavioral Characteristics

To investigate the psychological causes of public panic and fear, this study examined ten factors contributing to COVID-19-related panic levels: multifaceted online information, post-recovery sequelae, mask-wearing prevalence, official notifications, public rumors, extensive media coverage, unclear etiology, high mortality rates, lack of effective treatments, COVID-19 lethality, and COVID-19 transmissibility. Analytical results are presented in Table 8.

**Table 8.** Results of t-test and LSD Analysis (Psychological Panic)

| I | J | I-J | p |
| --- | --- | --- | --- |
| Sex |  |  |  |
| Male | Female | -0.03052* | 0.007 |
| Age |  |  |  |
| <20 | 20-29 | -0.31579* | 0.002 |
|  | 30-39 | -0.35203* | <0.001 |
|  | 40-49 | -0.29440* | 0.017 |
|  | ≥50 | -0.29808 | 0.053 |
| Occupation |  |  |  |
| Corporate Employee | Student | 0.09701* | 0.008 |

Independent samples t-test revealed significantly higher panic levels among females compared to males (p *<* 0.01). Social role theory posits that women traditionally assume familial health-monitoring roles, internalizing a hypersensitive monitoring tendency toward disease threats. During the pandemic, women not only assessed personal infection risks but also conducted proxy risk assessments for family members (particularly children and elders), a dual responsibility exacerbating psychological stress. Thus, heightened attention to female emotional responses is warranted.

LSD method analysis indicated the lowest panic impact among individuals under 20 years old (p *<* 0.06). Panic often stems from exaggerated imaginings of unknown threats; younger populations exhibit limited comprehension of complex viral information (e.g., lethality, transmission mechanisms), resulting in lower risk perception capacity and reduced overreaction. Additionally, adolescents prioritize immediate emotional needs (e.g., academic pressure, social interactions) over long-term health consequences (e.g., sequelae), necessitating targeted guidance.

Occupational comparisons showed significantly higher panic effects among corporate employees versus students (p *<* 0.01). Corporate workers face occupational uncertainty and income instability, with pandemic-induced business closures directly threatening survival needs (e.g., unemployment, salary cuts). Chronic workplace stress also predisposes them to physiological issues (e.g., insomnia, immune dysfunction), amplifying psychological distress. Conversely, students received systemic protections (e.g., online education, mental health support) and relied on familial financial security, mitigating direct survival pressures.

Further analysis of the ten primary sources of pandemic-related panic (Table 9) identified the following as dominant contributors to public fear and anxiety: post-recovery sequelae (mean = 4.34), multifaceted online information (4.20), unclear etiology (4.19), and mask-wearing compliance coupled with official notifications (4.06). Less impactful factors included extensive media coverage (3.91), rapid lethality of the virus (3.87), public rumors (3.66), lack of effective treatments (3.59), viral transmissibility (3.43), and high patient mortality rates (3.21).

**Table 9.** Public Psychological Panic Sources Regarding COVID-19.

| Panic Sources | Mean Impact Score | Panic Sources | Mean Impact Score |
| --- | --- | --- | --- |
| 1.Lack of effective treatments | 3.59 | 6. Extensive media coverage | 3.91 |
| 2.Rapid lethality of the virus | 3.87 | 7. Multifaceted online information | 4.20 |
| 3.COVID-19 transmissibility | 3.43 | 8. Mask-wearing prevalence & official notifications | 4.06 |
| 4.High patient mortality rates | 3.21 | 9. Unclear etiology | 4.19 |
| 5.Public rumors and fears | 3.66 | 10. Post-recovery sequelae | 4.34 |

In 2022, the COVID-19 resurgence was mainly driven by the Omicron variant, which, compared to earlier Delta variant, has the characteristics of “high transmissibility and low fatality”. According to the China CDC Weekly data as of May 3, 2022, the overall severe - case rate among Omicron - infected individuals was only 0.065% (22/33816), mostly in elderly patients with underlying diseases. Also, the “risk amplification effect” due to information asymmetry in the pandemic’s early stage, such as excessive panic and hospital overcrowding, eased in 2022. The public, through multiple pandemic waves, gained response experience and became more familiar with prevention and control measures (like home quarantine and nucleic acid testing), reducing anxiety and fear from uncertainty.

Additionally, data from Table 10 indicate that public psychological states remained relatively stable. Among the twelve items, six were positive indicators: generally happy (3.40), life is interesting (3.42), bravely facing problems (3.89), concentrating on tasks (3.79), making independent decisions (3.75), and playing important roles in various aspects (3.57). The remaining six were negative indicators: depressed and unhappy mood (2.60), experiencing psychological stress (2.80), “unable to overcome difficulties (2.25), loss of confidence (2.10), feeling worthless (1.91), and insomnia due to worries (2.27). Positive indicators scored higher, while negative indicators scored lower. In summary, public psychological well-being remained relatively intact; although the pandemic impacted public mentality, the severity was limited.

**Table 10.** Public Psychological States During COVID-19.

| Psychological State | Frequency | Psychological State | Frequency |
| --- | --- | --- | --- |
| 1. Generally happy | 3.40 | 7. Maintaining concentration | 3.79 |
| 2. Feeling depressed | 2.60 | 8. Loss of confidence | 2.10 |
| 3. Finding life interesting | 3.42 | 9. Making independent decisions | 3.75 |
| 4. Experiencing psychological stress | 2.80 | 10. Feeling worthless | 1.91 |
| 5. Bravely facing problems | 3.89 | 11. Playing useful roles in multiple areas | 3.57 |
| 6. Unable to overcome difficulties | 2.25 | 12. Insomnia due to worries | 2.27 |

This study further analyzed public psychological tension during the pandemic (Table 11), revealing the lowest tension levels among service industry workers (3.07). This is attributed to their prolonged exposure to high-frequency interpersonal interactions (e.g., catering, retail, logistics), which fosters adaptive responses to potential risks. Additionally, certain service roles (e.g., delivery riders, ride-hailing drivers) involve significant job autonomy, enabling risk mitigation through workload adjustments or low-risk service area selection. Conservation of Resources theory suggests such perceived control effectively alleviates anxiety. Healthcare workers also exhibited relatively low psychological tension (3.12). Occupational exposure adaptation in medical professionals triggers psychological immunization—repeated exposure to biological stressors induces emotional desensitization. Moreover, teamwork and institutional support (e.g., guaranteed protective equipment) enhance collective efficacy, offsetting individual vulnerability perceptions and maintaining emotional homeostasis in high-risk settings. Civil servants demonstrated stable psychological states (3.13), likely linked to their occupational security.

**Table 11.** Public Psychological Tension During COVID-19.

| Occupation | M | Occupation | M |
| --- | --- | --- | --- |
| Civil Servant | 3.13 | Self-employed | 3.21 |
| Corporate Employee | 3.19 | Student | 3.23 |
| Service Worker | 3.07 | Professional in S/E/C | 3.17 |
| Healthcare Worker | 3.12 | Farmer | 4.00 |
| Laborer | 3.30 | Other | 3.14 |

Among all negative indicators, rural residents exhibited significantly higher pandemic-related anxiety compared to other groups. This disparity stems from two interrelated factors: rural areas face underdeveloped medical infrastructure, with the number of hospital beds per 1,000 people being only 36% of urban levels (National Health Commission, 2020), coupled with insufficient critical care capacity and drug reserves, exacerbating anticipatory anxiety. Simultaneously, rural households rely heavily on migrant work income (62.3% of disposable income; National Bureau of Statistics, 2020). Pandemic-induced disruptions to employment and agricultural product oversupply have precipitated livelihood crises, amplifying economic and psychological pressures that intensify pandemic concerns.

### 4.4 Analysis of Public Emergency Response Behaviors

Psychological research on risk perception demonstrates that unfamiliar or uncontrollable information during risk events is the primary driver of psychological tension. The findings of this questionnaire survey align with this conclusion. Public concerns about COVID-19’s etiology, treatment uncertainties, and unpredictability remain key contributors to panic. This study argues that targeted media strategies—emphasizing transparency, timeliness, and alignment with public information priorities—can mitigate anxiety. Excessive dissemination of unprocessed negative information may amplify panic; thus, information providers must assist the public in filtering credible content to enhance crisis preparedness.

Public risk perception significantly predicts proactive behavioral adaptations. Higher familiarity with pandemic risks correlates with increased adoption of adaptive behaviors. When continuously exposed to risks, individuals typically engage in actions to alleviate stress. Improved objective understanding of the pandemic promotes behaviors such as frequent hand disinfection, reduced social contact, consistent mask usage, health knowledge dissemination, and emotional coping strategies (e.g., stress eating), thereby reducing anxiety. These observations are consistent with existing research.

As shown in Table 12, public hygiene habits improved significantly during the pandemic, such as frequent hand disinfection (4.15), reduced contact in public spaces (3.90), and consistent mask usage when going out (4.42). These findings indicate not only widespread adoption of preventive measures but also the effectiveness of governmental health communication campaigns in China. Government-issued information scientifically guided public behavior, enhancing COVID-19 prevention capabilities and confidence.

**Table 12.** Public Behavioral Changes During COVID-19.

| Behavioral Change | Frequency | Behavioral Change | Frequency |
| --- | --- | --- | --- |
| 1. Frequent hand disinfection | 4.15 | 5. Increased workload | 2.63 |
| 2. Reduced social contact | 3.90 | 6. Insufficient or poor-quality sleep | 2.49 |
| 3. Consistent mask usage when going out | 4.42 | 7. Overeating to relieve stress | 2.03 |
| 4. Discussing and promoting preventive knowledge | 3.52 | 8. Praying to ancestors or deities for protection | 2.18 |

Table 13 demonstrates that LSD method analysis revealed individuals with bachelor’s degrees or higher (high education group) adopted more science-based COVID-19 prevention strategies compared to those with associate degrees or below (low education group). The high education group scored significantly higher in practices like hand disinfection (p *<* 0.06). Highly educated individuals typically possess accurate epidemiological understanding and prioritize evidence-based medical advice (e.g., masking, vaccination) over empirical or folk remedies, leading to more scientific preventive behaviors.

**Table 13.** Results of t-test and LSD Analysis (Coping Behaviors)

| I | J | I-J | p |
| --- | --- | --- | --- |
| Education(proactive) |  |  |  |
| ≥ Bachelor | ≤ Associate | 0.08536 | 0.058 |
| Sex(avoidant) |  |  |  |
| Male | Female | -0.04895 | 0.099 |

Additionally, t-tests indicated that females were more likely than males to adopt passive coping behaviors (p *<* 0.1). This aligns with prior findings: women exhibit heightened vigilance toward family health risks (e.g., elderly and children), with studies showing their concern intensity for familial infection risks is 1.3× higher than males. This excessive sense of responsibility predisposes them to chronic anxiety, triggering passive coping mechanisms.

### 4.5 Correlation Analysis Among Variables

The above analyses demonstrate that both positive and negative pandemic risk information significantly influence public risk perception, triggering either panic-driven negative mindsets or adaptive positive attitudes, which subsequently drive distinct coping behaviors. However, due to evolving viral characteristics, socio-psychological adaptation, dynamic information dissemination mechanisms, and adjustments in social support systems, public risk perception during the 2022 pandemic phase differed from earlier stages. Building on prior research by Shi Kan et al. (2020), this study constructed a theoretical model framework with risk perception as a mediating variable to compare the differential impacts of pandemic information on risk cognition and coping behaviors across two COVID-19 outbreak periods, thereby refining the understanding of underlying mechanisms. Specifically, Model 1 was designed as follows: four categories of pandemic-related information served as independent variables; proactive and avoidant coping behaviors as dependent variables; risk perception as the mediator; and psychological tension as a moderating variable regulating the mediation effect. Pearson correlation tests were conducted among these variables, yielding the following empirical results:

As shown in Table 14^3^, recovery-related information exhibited a significant positive correlation only with proactive coping behaviors. In contrast, infection-related information, personally relevant information, and government preventive measures correlated positively with both proactive and avoidant coping behaviors. Strong positive correlations were observed between infection-related information and government measures (r = 0.615) and between personally relevant information and government measures (r = 0.561), suggesting that increased negative pandemic information may prompt amplified governmental interventions. Psychological tension showed a moderately strong positive correlation with avoidant coping (r = 0.339), indicating that heightened tension likely exacerbates passive responses. Additionally, public risk perception positively correlated with government measures (r = 0.214), underscoring the public’s focus on official containment strategies.

**Table 14.** Correlation Analysis Among Variables.

|  | 1 | 2 | 3 | 4 | 5 | 6 | 7 |
| --- | --- | --- | --- | --- | --- | --- | --- |
| 1 | 1 |  |  |  |  |  |  |
| 2 | 0.538** | 1 |  |  |  |  |  |
| 3 | 0.557** | 0.413** | 1 |  |  |  |  |
| 4 | 0.615** | 0.454** | 0.561** | 1 |  |  |  |
| 5 | 0.125** | 0.237** | 0.139** | 0.214** | 1 |  |  |
| 6 | 0.139** | 0.219** | 0.107** | 0.171** | 0.535** | 1 |  |
| 7 | 0.073** | -0.047 | 0.083** | 0.079** | -0.364** | -0.669** | 1 |

Note: \* indicates relatively significant differences; \*\* indicates significant differences.

These findings imply that enhancing the transparency of governmental prevention campaigns and disseminating positive information (e.g., updates on antiviral drug development) could mitigate public panic and psychological tension while fostering proactive attitudes toward pandemic management.

Building on this foundation, the study tested the previously constructed Model 1. After adjusting the model settings based on modification indices, the goodness-of-fit indices are presented in Table 15.

**Table 15.** Goodness-of-Fit Indices of the Model.

| Model | $\chi^2$ | df | GFI | AGFI | CFI | TLI | RMSEA |
| --- | --- | --- | --- | --- | --- | --- | --- |
| 1 | 16985.387 | 1529 | 0.519 | 0.480 | 0.673 | 0.659 | 0.079 |
| 2 | 10384.854 | 1526 | 0.713 | 0.689 | 0.813 | 0.804 | 0.060 |
| 3 | 5285.866 | 1523 | 0.883 | 0.873 | 0.920 | 0.917 | 0.039 |

Model 2 expanded upon the initial hypothesized Model 1 by adding two paths: one from infection-related information and another from personally relevant information (both negative pandemic indicators) to avoidant coping behaviors. These additions significantly improved all model fit indices. Subsequently, Model 3 further incorporated a path from government preventive measures (a positive pandemic indicator) to proactive coping behaviors. The final Model 3 demonstrated robust goodness-of-fit indices: all metrics exceeded 0.85, with CFI and TLI indices surpassing 0.9 and RMSEA below 0.5, confirming its theoretical validity and optimal fit. Consequently, Model 3 was finalized as the definitive public risk perception model, as illustrated in Figure 4.

**Fig 4.**
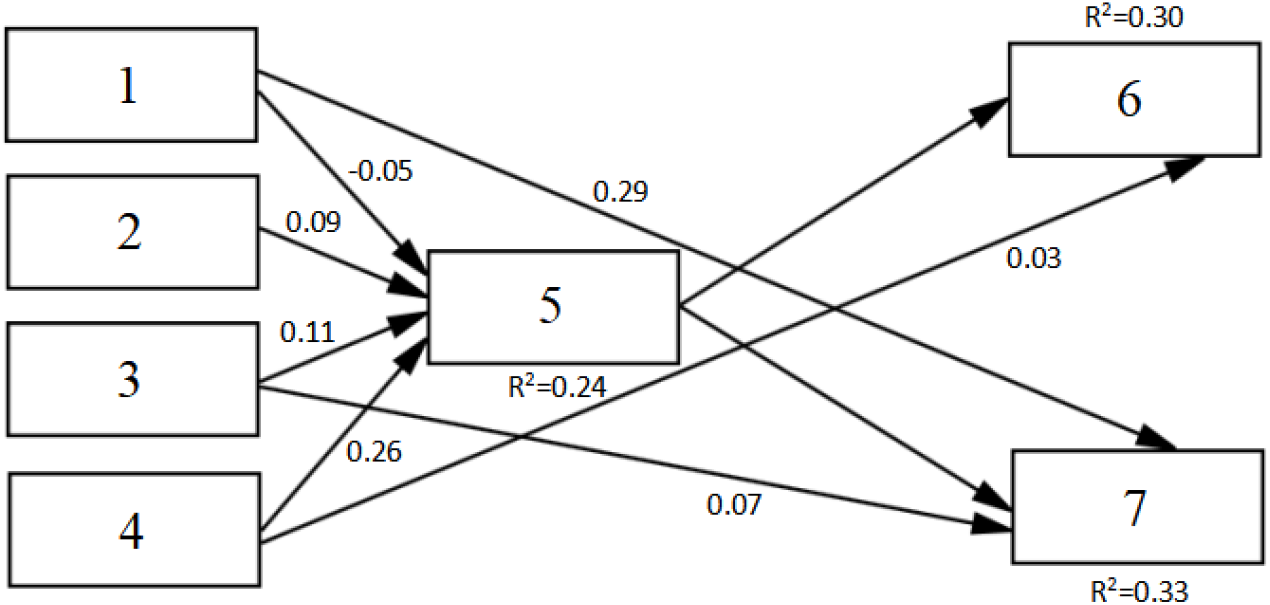
Public Risk Perception and Psychological Behavior Prediction Model

As illustrated in Figure 4, distinct pandemic information factors differentially impact public risk perception. Infection-related information exerts a negative influence on risk perception. Prolonged exposure to infection data (e.g., daily case updates) normalizes COVID-19 as a routine health threat rather than a survival crisis, thereby attenuating risk sensitivity. In contrast, recovery-related information and government preventive measures positively correlate with risk perception (positive path coefficients), aligning with Shi Kan et al.’s 2020 findings. Notably, personally relevant information also demonstrated a positive influence on risk perception in this study. This is because, in the initial stage of the COVID - 19 pandemic, the high pathogenicity and fatality rate of the coronavirus (the original strain and the Delta variant) triggered public panic to a great extent.Amid vaccine/therapeutic uncertainties, proximity to confirmed cases became a critical risk reference. Absence of local infections fostered perceptions of controllable risk, lowering overall risk perception. But now, the high transmissibility of the Omicron variant has caused a surge in the number of infected people. surging case numbers—amplified via social media and community networks—triggered exposure risk alertness, compounded by viral mutation uncertainties, thus heightening risk perception. All path coefficients reached statistical significance.

Personally relevant information uniquely functions as a dual-path influencer: it indirectly promotes proactive coping behaviors by elevating risk perception while directly increasing avoidant coping behaviors, thereby validating Hypothesis 5.

Additionally, both positive and negative pandemic information positively affect risk perception, confirming Hypothesis 3.

The model integrates proactive/avoidant coping behaviors and mental health indicators, demonstrating that COVID-19 information impacts psychological behaviors through the mediating role of risk perception. Subsequent mediation and moderation analyses (Table 16) focused on infection-related information as the independent variable, with other variables yielding consistent results.

**Table 16.** Mediation and Moderation Effects Tests.

| Variables | Equation 1<br>(Proactive Coping) |  |  |  | Equation 2<br>(Risk Perception) |  |  |  | Equation 3<br>(Proactive Coping) |  |  |  |
| --- | --- | --- | --- | --- | --- | --- | --- | --- | --- | --- | --- | --- |
| | $\beta$ | SE | t | 95%CI | $\beta$ | SE | t | 95%CI | $\beta$ | SE | t | 95%CI |
| Sex | 0.01 | 0.02 | 0.68 | [-0.03, 0.06] | -0.11 | 0.02 | -4.39 | [-0.16, -0.06] | 0.01 | 0.02 | 0.68 | [-0.03, 0.06] |
| Age | 0.15 | 0.02 | 7.20 | [0.08, 0.14] | 0.11 | 0.02 | 6.40 | [0.08, 0.14] | 0.11 | 0.01 | 7.20 | [0.08, 0.14] |
| Education | -0.05 | 0.02 | -2.53 | [-0.08, -0.01] | 0.08 | 0.02 | 4.00 | [0.04, 0.12] | -0.04 | 0.02 | -2.53 | [-0.08, -0.01] |
| Infection | 0.07 | 0.01 | 3.23 | [0.02, 0.07] | 0.07 | 0.02 | 4.29 | [0.04, 0.12] | 0.04 | 0.01 | 3.23 | [0.02, 0.07] |
| Risk | 0.51 | 0.02 | 23.74 | [0.46, 0.54] |  |  |  |  | 0.50 | 0.02 | 23.73 | [0.45, 0.54] |
| Tension |  |  |  |  | 0.07 | 0.09 | 0.78 | [-0.10, 0.24] |  |  |  |  |
| Interaction |  |  |  |  | -0.03 | 0.02 | -1.33 | [-0.08, 0.01] |  |  |  |  |
| R<sup>2</sup> |  |  | 0.32 |  |  |  | 0.07 |  |  |  | 0.32 |  |
| F |  |  | 148.78*** |  |  |  | 19.45*** |  |  |  | 148.78*** |  |

As shown in Table 16, after controlling for sex, age, and educational attainment, the regression coefficient of infection-related information on risk perception was significant (*β* = 0.07, p *<* 0.05). After controlling for infection-related information, the regression coefficient of risk perception on proactive coping was significant (*β* = 0.50, p *<* 0.001). Bootstrapping results confirmed a significant indirect effect: the 95% confidence interval [0.45, 0.54] excluded zero, validating the mediation effect. Shi Kan et al.’s 2020 study demonstrated that, after controlling for sex, age, and education, the interaction term between recovery-related information and psychological tension significantly predicted risk perception (*β* = 0.06, p *<* 0.05), supporting moderation effects. However, this study found the interaction term’s regression coefficient on risk perception to be non-significant (p *>* 0.05) under similar controls, contradicting prior findings. This discrepancy arises because heightened panic and tension during initial exposure to public health risks impair rational decision-making, distorting risk perception accuracy and efficacy [27]. So, the study posits that psychological tension amplifies risk perception during initial encounters with novel threats. By the 2022 outbreak phase, however, repeated risk exposure desensitized public psychological defenses, attenuating emotional reactivity to risk stimuli. Additionally, the 2020 information vacuum forced reliance on subjective psychological states for risk appraisal, whereas 2022’s information overload shifted public dependence to authoritative quantitative metrics (e.g., infection rates, severe case ratios), reducing the moderating weight of subjective psychological states. These dynamics validate Hypothesis 2.

## 5 Conclusions and Prospects

### 5.1 Conclusions

This study, situated within the context of the COVID-19 pandemic as a major public health emergency, conducted a nationwide questionnaire survey across 32 provinces in China to construct a predictive model of public socio-psychological behaviors centered on risk perception. The research aimed to dissect public risk cognition characteristics and behavioral responses during the pandemic, as well as compare differences in risk perception and coping strategies between the initial and secondary outbreak phases. Employing stratified sampling to ensure representation across regions, pandemic stages, and social demographics, 1,605 valid questionnaires were collected. Through reliability/validity tests, factor analysis, correlation analysis, and structural equation modeling (SEM), the following conclusions were drawn:

1. During the survey period, public risk perception of COVID-19 leaned toward familiarity and controllability. Compared to the 2020 risk cognition map by Shi Kan et al., dimensions such as “cure rates” and “post-recovery contagiousness” shifted from unfamiliar/uncontrollable to familiar/controllable, reflecting improved public risk cognition. However, “post-recovery health impacts” and “viral etiology” remained in intermediate controllability zones, representing persistent vulnerabilities that may amplify panic.
2. Government preventive measures exerted the strongest influence on public risk perception, significantly surpassing the effects of infection-related information, recovery-related data, and personally relevant information.
3. Public risk awareness and fear levels notably decreased compared to the pandemic’s initial phase.
4. Negative pandemic information (e.g., infection statistics) reduced risk perception, while positive information (e.g., recovery updates, government measures) enhanced it. Notably, “personally relevant information,” despite its negative valence, positively influenced risk perception.
5. SEM results confirmed that COVID-19 impacts psychological behaviors through the mediating role of risk perception, with predictive indicators including psychological tension, coping behaviors, and mental health.
6. “Personally relevant information” uniquely functioned as a dual-path driver: it indirectly promoted proactive coping via heightened risk perception while directly increasing avoidant coping.
7. Moderation tests revealed non-significant effects of interaction terms on risk perception (p *>* 0.05), contrasting with Shi Kan et al.’s 2020 findings. This suggests dynamic adaptability in public socio-psychological and behavioral patterns during protracted health crises.

### 5.2 Limitations and Future Directions

Despite its contributions, this study has limitations. First, while covering 32 provinces, the sample size remains limited, and potential sampling biases may constrain representativeness. Second, reliance on self-reported questionnaires risks subjective biases and data incompleteness. Third, longitudinal data collection across pandemic phases (escalation, peak, decline) introduced non-response biases due to temporal variations in respondent characteristics. Fourth, analytical methods, though robust, may lack precision in capturing complex psychological-behavioral dynamics.

Future research should expand sample sizes, employ mixed-method approaches (e.g., interviews, observational data), and integrate advanced analytics to holistically unravel risk cognition and coping mechanisms in crises. Longitudinal comparisons of pandemic waves across different timelines could reveal temporal patterns, while cross-cultural and socioeconomic analyses would enhance the generalizability of findings. Such efforts will strengthen theoretical foundations for evidence-based emergency policymaking.

## Author Contributions

Conceptualization,Ge Shi,Liyi Ma;methodology,Ge Shi,Liyi Ma,Xingtong Men,Jiayi Li, and Chenchen Xia;formal analysis,Liyi Ma,Xingtong Men,Jiayi Li, and Chengzi Huang;investigation,Xingtong Men,Jiayi Li,Chengzi Huang,andChenchen Xia;data curation,Ge Shi,Liyi Ma;writing-original draft preparation,Ge Shi,Liyi Ma,Xingtong Men,Jiayi Li,and Chengzi Huang;writing-review and editing,Ge Shi,Liyi Ma,Xingtong Men,Jiayi Li,and Chengzi Huang;resources,Ge Shi,Liyi Ma;project administration,Ge Shi,Liyi Ma.All authors have read and agreed to the published version of the manuscript.

## Data Availability Statement

All data necessary to replicate our study’s findings are publicly available in https://figshare.com/, with the dataset assigned the following DOI: 10.6084/m9.figshare.28696247.

## Footnotes

1 China’s National Health Commission announced on December 25, 2022, that daily pandemic updates would no longer be published.

2 The numbering system employed in Figures 2 and 3 maintains referential consistency with Table 7.

3 For clarity and conciseness, the table displays numeric codes only. The codes correspond to: 1 = infection-related information, 2 = recovery-related information, 3 = personally relevant information, 4 = government preventive measures, 5 = risk perception, 6 = proactive coping, 7 = avoidant coping. The model diagram (Figure 4) follows the same conventions.

## References

1. Adhanom T. WHO Director-General’s opening remarks at the media briefing on COVID-19; 2020. Available from: https://www.who.int/director-general/speeches/detail/who-director-general-s-opening-remarks-at-the-media-briefing-on-covid-1911-march-2020.

2. Jing M. Risk Perception and Behavioral Threshold: Characteristics, Causes and Prevention Strategies of Communication Risks. Journal of Northwest Normal University (Social Sciences Edition). 2023;60(3):96–104.

3. Wang J, Zhang Y. Risk Cognition, Social Sentiment, and Future Expectations: Changes in Social Mentality Across Different Pandemic Stages. Social Science Front. 2022;(10):220–237.

4. Müller SR, Delahunty F, Matz SC. The impact of the early stages of COVID-19 on mental health in the United States, Germany, and the United Kingdom. Journal of Personality and Social Psychology. 2023;124(3):620.

5. Cori L, Bianchi F, Cadum E, Anthonj C. Risk perception and COVID-19; 2020.

6. Xu J, Xue L, Shou M. Public Risk Perception in Environmental Governance: A Half-Century Review and Future Prospects. China Public Administration Review. 2016;13(2):87–105.

7. Ge M. Risk Framing, Amplification and Governance of Rumors in Public Health Events: An Analysis of Early COVID-19 Online Rumors. Journalism Research. 2023;(8):44–58, 118.

8. Xia J, Meng Y, Wen F, Li H, Meng K, Zhang L. Caring for anxiety among adults in the face of COVID-19: A cross-sectional online survey. Journal of Affective Disorders Reports. 2020;1:100014.

9. Zhao H, Ge Y, Yin C. How Pandemic Information Affects Pandemic Evolution: A Computational Experimental Model. Global Media Journal (China Edition). 2024;46(5):133–159.

10. Garfin DR, Silver RC, Holman EA. The novel coronavirus (COVID-2019) outbreak: Amplification of public health consequences by media exposure. Health psychology. 2020;39(5):355.

11. Kim L, Fast SM, Markuzon N. Incorporating media data into a model of infectious disease transmission. PloS one. 2019;14(2):e0197646.

12. Arafat SY, Islam MA, Kar SK. Mass media and panic buying. In: Panic buying: Perspectives and prevention. Springer; 2021. p. 65–80.

13. Arafat SY, Kar SK, Menon V, Kaliamoorthy C, Mukherjee S, Alradie-Mohamed A, et al. Panic buying: An insight from the content analysis of media reports during COVID-19 pandemic. Neurology, Psychiatry and Brain Research. 2020;37:100–103.

14. Cohen J, Kupferschmidt K. Countries test tactics in ‘war’against COVID-19; 2020.

15. Lee J, Ahn Y. Risk Perception and Behavioral Changes During the COVID-19 Pandemic: A Meta-Analytic Review. Journal of Public Health Psychology. 2023; p. 1–15. doi:10.13222/jph.2023.0011.

16. Simon HA. Rationality in psychology and economics. Journal of business. 1986; p. S209–S224.

17. Kahneman D, Slovic P, Tversky A. Judgment under uncertainty: Heuristics and biases. Cambridge university press; 1982.

18. Slovic P. Perception of risk. science. 1987;236(4799):280–285.

19. Jiao S, Shi K, Zhou H, Guo H, Gao W. Public Psychological States and Emotional Guidance Strategies Under COVID-19 Risk Information Exposure. Medicine and Society. 2020;33(5):98–104.

20. Shi K, Lu J, Fan H, Jia J, Song Z, Li W, et al. Rationality Characteristics and Psychobehavioral Prediction Model of the Public in 17 Cities During SARS Crisis. Chinese Science Bulletin. 2003;(13):1378–1383.

21. Institute of Psychology, Chinese Academy of Sciences. Healthy Mentality: Overcoming SARS through Psychological Coping. Beijing: Science Press; 2003.

22. Powell D. An Introduction to Risk Communication and the Perception of Risk. Financial Analysts Journal (March-April. 2004;.

23. Moss R. Coping Responses Inventory-Adult Form Professional Manual. Psychological Assessment Resources: Lutz, FL, USA. 1993;.

24. Wen F, Shi K, Zhou H, Guo H, Gao W. “Ripple Effect” and “Psychological Typhoon Eye Effect”: A Dual-Perspective Examination of Risk Perception and Anxiety in COVID-19 Pandemic Areas. Acta Psychologica Sinica. 2020;52(9):1121–1140.

25. Wang J, editor. Annual Report on Social Mentality in China (2020). Beijing: Social Sciences Academic Press; 2021.

26. Wu L. “Information Immunity” in Major Public Health Emergencies. People’s Tribune. 2020;(S1):116–119.

27. Geng S. Impact of COVID-19 Pandemic Risk Perception on Post-Traumatic Stress Disorder Among Healthcare Workers. Southern Medical University. Guangzhou, China; 2022.

